# Laser Coronary Atherectomy and polymeric coronary wires in uncrossable lesions – A word of caution

**DOI:** 10.1101/2023.06.21.23291723

**Authors:** Giorgio Marengo, Carlos Cortés, Juan Pablo Sánchez-Luna, Jose Carlos Gonzalez-Gutiérrez, Javier Gómez Herrero, Jorge Sanz-Sanchez, Hipólito Gutiérrez, Ana Serrador, Alberto Campo, Sara Blasco-Turrión, Gabriele Gasparini, J. Alberto San Román, Ignacio J. Amat-Santos

## Abstract

**Background:** Excimer laser coronary atherectomy (ELCA) represents one of the last available options in case of balloon uncrossable lesions but no bench testing or clinical experience has been reported with polymeric wires. We aimed to assess whether ECLA use can disrupt or melt the cover and coating of coronary wires.

**Methods:** A total of 22 wires (10 polymer-jacketed and 12 hydrophilic non polymeric) were tested in an uncrossable lesion model. Two ELCA tests were performed over each wire at 8 and 4 cms from the tip with low and high settings, respectively. Microscopic analysis was performed at baseline images and after each test. Wire disruption was classified as Grade 1 (superficial scratches), Grade 2 (coil damage or solution of continuity of the polymeric cover) and Grade 3 (wire rupture/core disruption or de-coiling).

**Results:** After 44 ELCA simulations, wire disruption occurred in 16 cases (36.3%). Overall, events were more common for polymer-jacketed than for hydrophilic wires (12 vs 4, p=0.004). No grade 3 events occurred. Grade 2 events occurred in 9 cases (20.5%) and were more frequent with polymer- jacketed wires (8 vs 1, p=0.006). With low ELCA settings only polymer-jacketed wires suffered disruption (5 vs 0, p=0.009). With higher settings 11 events occurred (7 of Grade 2 and 4 of Grade 1) and incidence of grade 2 events was higher for polymer-jacketed wires (6 vs 1, p=0.02).

**Conclusions:** ELCA might be a safe option for coronary uncrossable lesions but its use with high settings over polymer-jacketed wires is discouraged.

**CONDENSED ABSTRACT:** We aimed to assess whether ECLA use can disrupt coronary wires in balloon/microcatheter uncrossable lesion model. Forty-four in-vitro experiments were performed over hydrophilic-coated and polymer-jacketed coronary wires with different ELCA settings. Wire disruption was classified as Grade 1 (scratches), Grade 2 (coil or polymeric cover damage) and Grade 3 (core disruption/de- coiling). Wire disruption occurred 36.3% of cases and was more common for polymer-jacketed wires than for hydrophilic-coated wires (p=0.004). With high ELCA settings the incidence of grade 2 events was higher for polymer-jacketed wires (p=0.02). This discourages the use of ELCA with high settings over polymer-jacketed wires.

## INTRODUCTION

Coronary lesions are defined balloon-uncrossable when they cannot be crossed with any balloon despite a successful distal crossing with a coronary wire. [1,2] They are encountered in up to 9% of chronic total occlusions (CTO) and can be also found in non-occlusive coronary lesions. [3,4] Multiple techniques have been described to face uncrossable lesions, aiming to modify the plaque or to increase the support. Multi-step algorithms have been designed, starting with support improvement and with “safer” small-balloon techniques and ending with more difficult and risky approaches, such as subintimal techniques. [5–7]

In this context, rotational atherectomy has been described as a safe and effective tool, but it is often doomed by the need of wire exchange through a microcatheter, that can be extremely challenging in uncrossable lesions. Various approaches have been proposed to provide a successful advancement of low-profile coronary tools such as small balloons or microcatheters and nowadays one of the most often chosen is the excimer laser coronary atherectomy (ELCA). [8–10] ELCA uses a xenon-chloride monochromatic exciter laser to produce ultraviolet light that is absorbed by the plaque, releasing a pressure wave that leads to plaque vaporizing and debulking. [11] According to the LAVA registry [12] the most common indication for its use in the USA are balloon-uncrossable lesions (43.8% of patients), with a procedural success over 90% and a low incidence of complications.

Despite the high success rate [13–17], some concerns have been raised about the potential risk of damage of the wire coating, especially when a prolonged laser pulsing with high fluence and frequency is performed or when contrast is injected during laser pulsing to increase the photomechanical effect of ELCA. [18] Nowadays, no specification regarding recommended wire type is provided in the instructions for use and often, ELCA is performed over a stiffer polymer- jacketed wire, especially when a microcatheter cannot be advanced to exchange the wire. In our clinical practice wire disruption with ELCA after successful distal wiring with a polymeric wire has been detected as shown in **Supplementary Fig 1**. For this reason, we performed an experimental study aiming to assess the safety and the impact of the coronary laser over the structure of different hydrophilic-coated and polymer-jacketed wires.

## METHODS

### Study Design

#### In vitro model

A plastic model of a coronary artery filled with saline solution was used (Supplementary fig 2). The uncrossable lesion, where the coronary laser was stuck, was empirically reproduced at 8 cm (7.5-8.5 cm) at a 4 cm (3.5-4.5 cm) from the tip of the wire. No mechanical stress (pushing) was applied over ELCA catheter, but position was fixed.

#### Coronary Laser

The CVX-300 coronary excimer laser system with 0.9 mm caliber made by Spectranetics (Colorado Springs, CO, USA, acquired by Phillips) was used. To simulate the mechanism of a balloon-uncrossable lesion, the laser was advanced over the wire and was stuck where the uncrossable lesion was simulated. After laser calibration, seven pulses of ELCA were sequentially applied over each wire at 8 cm (pulse frequency of 25 pulses/sec and pulse fluence of 45 mj/mm2) and at 4 cm (pulse frequency of 40 pulses/sec and pulse fluence of 60 mj/mm2) from the tip of the wire (**Supplementary fig 2**).

#### Microscope

The DM202 MAX microscope made by Tomlov was used. We stored images of each wire at magnification power settings 10X and 15X before and after the ELCA. When we observed some noticeable effects of the ELCA over the wire, we further took other pictures at the operator’s discretion.

#### Wire selection

Coronary wires were divided in two groups according to the cover and coating characteristics: a) polymer-jacketed wires were defined when a polymer sleeve cover was fitted over a conventional spring-coil chassis, b) hydrophilic wires were defined when a hydrophilic coating was laid over the spring-coil. We perform test with coronary laser over 22 different coronary wires, either with hydrophilic coating or with an additional polymeric cover. We chose for our analysis both common workhorse wires and the most used specific CTO ones. Among the hydrophilic wires we included: Balance Middleweight (Abbott), Turntrac (Abbott), Powerturn Flex (Abbott), Infiltrac Plus (Abbott), Samurai (Boston Scientific), Judo 6 (Boston Scientific), Hornet 14 (Boston Scientific), Sion Blue (Asahi), Gaia 3 (Asahi), Confianza pro 12 (Asahi), Warrior (Teleflex) and Runthrough Extrafloppy (Terumo). Among the polymer-jacketed wires we included: Sion Black (Asahi), Fielder XT-A (Asahi), Fielder XT-R (Asahi), Gladius EX (Asahi), Gladius MG (Asahi), Whisper MS (Boston), Bandit (Teleflex), Raider (Teleflex), Progress 80 (Abbott), Pilot 200 (Abbott).

Before the simulation, 2 baseline images of each wire were stored (at 10X and 15X). Then, a test with ELCA (7 pulses) was performed over each wire at 8 cm and 4 cm from the wire tip (total of 44 tests). After the simulation each wire was re-assessed at the microscope, with the same settings and distances of the pre-test acquisitions.

#### Definition and classification of wire disruption

The magnitude of the wire disruption after ELCA was classified according to the following self-developed classification on structural damage of the wire (**Figure 1**): a) Grade 1: Superficial scratches of the more external layer of the wire, without an interruption in the integrity of the polymeric cover in case of polymer-jacketed wires; b) Grade 2: Damage of the wire coil or, in case of polymer-jacketed wires, when a solution of continuity of the polymeric cover (i.e., a hole or a cut) occurred. c) Grade 3: De-coiling, disruption of the wire core, or complete wire rupture.

**Figure 1:**
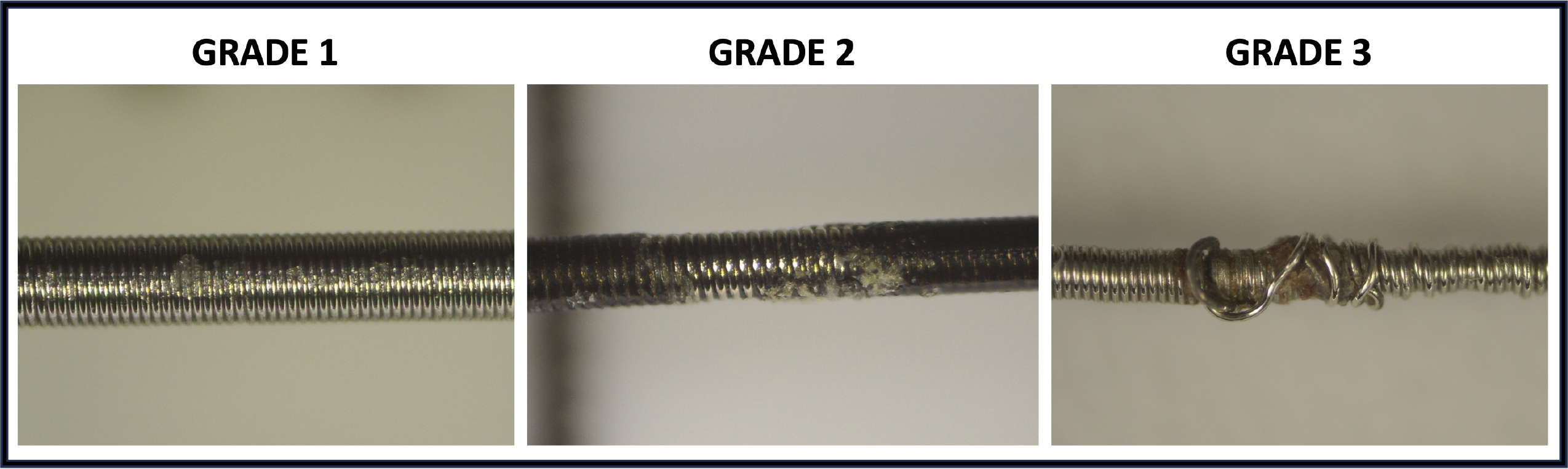
Classification of wire disruption. ***Legend:* (left panel)** Grade 1 wire disruption after ELCA simulation with superficial scratches of the external coil of a Runthrough Extrafloppy wire. **(central panel)** Grade 2 wire disruption after ELCA simulation with solution of continuity of the polymeric cover of a Fielder XT-A wire. **(right panel)** Grade 3 wire disruption with complete decoiling of the interested segment. No grade 3 events were observed in our simulation *(the wire shown in the image was not disrupted by coronary laser in our experiment)*. Abbreviations: ELCA: excimer laser coronary atherectomy.

#### Objectives

The primary objective was the detection of wire disruption and its degree after ELCA. The secondary objectives included the comparison of the incidence of wire disruption after ELCA according to covering type and to alternative frequency and fluence settings.

#### Statistical Analysis

Differences in the incidence of wire disruption between hydrophilic vs polymer-jacketed wires and after high vs. low ELCA settings were investigated by applying the exact Fisher’s test, considering 0.05 as significant threshold. Analyses were performed with SPSS® Statistics v24.

## RESULTS

ELCA tests were performed over 22 coronary wires. Among them 12 were hydrophilic non polymer-jacketed and 10 were polymer jacketed. A full list of the selected wires, with their specific features, is provided in **Table 1**. Two ELCA tests were performed over each wire (total of 44 tests) in 2 different segments, the first test was performed with lower settings (frequency of 25 pulses/sec and fluence of 45 mj/m2), and the second with higher settings (frequency of 40 pulses/sec and fluence of 60 mj/m2).

**Table 1:** characteristics of the included wires.

| NAME | COMPANY | TIP<br>LOAD | COATING | COVER | TYPE OF<br>POLYMER | DIAMETE<br>R |
| --- | --- | --- | --- | --- | --- | --- |
| SION BLUE | ASAHI | 0.5 g | Hydrophilic 18.5 cm<br>Hydrophobic<br>(Silicone distal tip 1.5 cm) | NO | / | 0.014" |
| GAIA 3 | ASAHI | 4.5 g | Uncoated distal tip<br>Hydrophilic 40 cm | NO | / | 0.014"<br>0.012" tip |
| CONQUEST PRO 12 | ASAHI | 12 g | Uncoated distal tip<br>Hydrophilic 20 cm | NO | / | 0.014"<br>0.009" tip |
| SION BLACK | ASAHI | 0.8 g | Hydrophilic 40 cm | YES<br>20 cm | Polyurethane<br>(no more information<br>given) | 0.014" |
| FIELDER XT-A | ASAHI | 1 g | Hydrophilic 17 cm | YES<br>17 cm | Polyurethane<br>(no more information<br>given) | 0.014"<br>0.010" tip |
| FIELDER XT-R | ASAHI | 0.6 g | Hydrophilic 17 cm | YES<br>17 cm | Polyurethane<br>(no more information<br>given) | 0.014"<br>0.010" tip |
| GLADIUS EX | ASAHI | 3 g | Hydrophilic 50 cm | YES<br>40 cm | Polyurethane<br>(no more information<br>given) | 0.014" |
| GLADIUS MG | ASAHI | 3 g | Hydrophilic 41 cm | YES<br>41 cm | Polyurethane<br>(no more information<br>given) | 0.014" |
| BALANCE<br>MIDDLEWEIGHT | ABBOTT | 0.6 | Hydrophilic | Intermediate<br>polymer | / | 0.014" |
| WHISPER MS | ABBOTT | 1 g | Hydrophilic | YES | Pellethane®<br>(aromatic<br>polyester based<br>thermoplastic<br>polyurethane) | 0.014" |
| TURNTRAC | ABBOTT | 0.8 | Hydrophilic | NO | / | 0.014" |
| INFILTRAC PLUS | ABBOTT | 13.9 g | Uncoated distal tip<br>Hydrophilic | NO | / | 0.014"<br>0.009" tip |
| PROGRESS 80 | ABBOTT | 11.5 g | Hydrophilic | YES | Pellethane®<br>(aromatic<br>polyester based<br>thermoplastic<br>polyurethane) | 0.014" |
| PILOT 200 | ABBOTT | 4.1 g | Hydrophilic | YES | Pellethane®<br>(aromatic<br>polyester based<br>thermoplastic<br>polyurethane) | 0.014" |
| POWERTURN<br>FLEX | ABBOTT | 1 g | Hydrophilic | NO | / |  |
| SAMURAI | BOSTON | 0.5 g | Reduced hydrophilic tip 1 cm<br>Hydrophilic 19 cm | NO | / | 0.014" |
| HORNET 14 | BOSTON | 14 g | Hydrophilic 15 cm | NO | / | 0.014"<br>0.008" tip |
| **JUDO 6** | BOSTON | 6 g | Hydrophilic 15 cm | NO | / | 0.014"<br>0.008" tip |
| **RUNTHROUGH**<br>**EXTRAFLOPPY** | TERUMO | 0.6 | Silicone distal tip 2 mm<br>Hydrophilic 24.8 cm | NO | / | 0.014" |
| **BANDIT** | TELEFLEX | 0.8 g | Hydrophilic 17 cm | YES<br>17 cm | Tecoflex®<br>(aliphatic<br>polyether-based<br>thermoplastic<br>polyurethane) | 0.014"<br>0.008" tip |
| **RAIDER** | TELEFLEX | 4 g | Hydrophilic 30 cm | YES<br>30 cm | Tecoflex®<br>(aliphatic<br>polyether-based<br>thermoplastic<br>polyurethane) | 0.014" |
| **WARRIOR** | TELEFLEX | 14 g | Hydrophilic 20 cm | NO | / | 0.014"<br>0.009" tip |

### Overall events

Overall incidence of wire disruption according to wire coating and to ELCA settings is shown in **Table 2**. After 44 ELCA simulations, 16 events of wire disruptions were registered (36.3%). Significantly more events occurred with polymer-jacketed wires than with hydrophilic- non-polymeric wires (12 vs 4, p = 0.004). According to ELCA settings, the overall incidence of wire disruption was higher with increased ELCA settings, but without reaching a statistical significance (11 vs 5 events, p = 0.11). No grade 3 disruptions were observed. Grade 2 disruptions were observed in 9 cases (20.5%), 7 with higher ELCA settings and 2 with low settings (p = 0.13). When polymer-jacketed wires were involved, grade 2 wire disruption was significantly more frequent (8 for polymeric vs 1 for hydrophilic, p = 0.006). Grade 1 disruptions were observed in 7 cases, without significant difference among polymer-jacketed wires and non-polymer-jacketed (4 events for polymer-jacketed wires vs 3 for hydrophilic wires, p = 0.68) and to ELCA settings (4 cases with higher ELCA settings and 3 with lower settings, p = 1).

**Table 2:** wire disruption events according to wire characteristics and to laser settings.

|  |  | WIRE CHARACTERISTICS |  |  | ELCA SETTINGS |  |  |
| --- | --- | --- | --- | --- | --- | --- | --- |
|  | Overall<br>(n = 44) | Polymer-jacketed<br>(n = 20) | Hydrophilic<br>(n = 24) | P value | High settings<br>(n = 22) | Low settings<br>(n = 22) | P value |
| Overall events | 16 (36.3%) | 12 (60%) | 4 (16.6%) | **0.004** | 11 (50%) | 5 (22.7%) | 0.11 |
| Grade 1 | 7 (15.9%) | 4 (20%) | 3 (12.5%) | 0.68 | 4 (18.1%) | 3 (13.6%) | 1 |
| Grade 2 | 9 (20.5%) | 8 (40%) | 1 (4.1%) | **0.006** | 7 (31.8%) | 2 (9%) | 0.13 |
| Grade 3 | 0 (0%) | 0 (0%) | 0 (0%) | / | 0 (0%) | 0 (0%) | / |

### ELCA low settings (25 pulses/sec x 45 mj/m2)

After the first ELCA simulation, with 7 pulses at 8 cm (7.5-8.5 cm) from the wire tip, with pulse frequency of 25 pulse/sec and pulse fluence of 45 mj/mm2, 5 wires suffered some grade of disruption (22.7%). All the events occurred amongst polymer-jacketed wires (5 vs 0, p = 0.009). No Grade 3 events occurred. Grade 2 events were observed in 2 cases (9%, with Sion Black and Fielder XT-A wire). Grade 1 events were observed in 3 cases (13%, with Fielder XT-R, Gladius EX and Bandit wire). **(Table 3).** No significant differences were observed comparing polymer-jacketed and non-polymer-jacketed wires for grade 2 and grade 1 disruptions with lower ELCA settings (respectively 2 vs 0, p = 0.19 and 3 vs 0, p = 0.07).

**Table 3:** wire disruption events according to wire characteristics after low and high settings ELCA simulation.

| *ELCA low settings (25 pulses/sec x 45 mj/mm2)* |  |  |  |  |
| --- | --- | --- | --- | --- |
|  |  | WIRE CHARACTERISTICS |  |  |
|  | Overall<br>(n = 22) | Polymer-jacketed<br>(n = 10) | Hydrophilic<br>(n = 12) | P value |
| **Overall events** | 5 (22.7%) | 5 (50%) | 0 (0%) | **0.009** |
| **Grade 1** | 3 (13.6%) | 3 (30%) | 0 (0%) | 0.07 |
| **Grade 2** | 2 (9%) | 2 (20%) | 0 (0%) | 0.19 |
| **Grade 3** | 0 (0%) | 0 (0%) | 0 (0%) | / |
| *ELCA high settings (40 pulses/sec x 60 mj/mm2)* |  |  |  |  |
|  |  | WIRE CHARACTERISTICS |  |  |
|  | Overall<br>(n = 22) | Polymer-jacketed<br>(n = 10) | Hydrophilic<br>(n = 12) | P value |
| **Overall events** | 11 (50%) | 7 (70%) | 4 (33.3%) | 0.19 |
| **Grade 1** | 4 (18.2%) | 1 (10%) | 3 (25%) | 0.59 |
| **Grade 2** | 7 (31.8%) | 6 (60%) | 1 (8.3%) | **0.02** |
| **Grade 3** | 0 (0%) | 0 (0%) | 0 (0%) | / |

For grading of wire disruption see Fig 1.

### ELCA high settings (40 pulses/sec x 60 mj/m2)

After the second ELCA simulation, with 7 pulses at 4 cm (3.5-4.5 cm) from the wire tip, with pulse frequency of 40 pulse/sec and pulse fluence of 60 mj/mm2, disruption occurred for 11 wires (50%). Again, polymer-jacketed wires were more frequently affected (7 events for polymer-jacketed vs 4 for hydrophilic wires) without reaching statistical significance (p = 0.19). No Grade 3 events occurred. Grade 2 event were observed in 7 cases (7/22 = 31.8%), both with polymer-jacketed wires (85.7%, with Bandit, Raider, Fielder XT-A, Fielder XT-R, Gladius MG and Sion Black wire) and with hydrophilic ones (14.3%, with Turntrac wire). Compared with hydrophilic wire, significantly more polymer-jacketed wires underwent grade 2 disruption with increased ELCA settings (6 vs 1, p = 0.02). Grade 1 events were observed in 4 cases (18,2%), both with hydrophilic wires (Gaia 3, Hornet 14, Runthrough Extrafloppy wires) and with polymer-jacketed ones (with Gladius EX wire), without significant differences according to the wire coating (3 vs 1, p = 0.59). All the wires that suffered any kind of disruption with the first ELCA experiment (Sion Black, Fielder XT-A, Fielder XT-R, Gladius EX and Bandit) underwent disruption also with the second ELCA test. **(Table 3).**

## DISCUSSION

When ELCA underwent pre-market bench testing and during its initial clinical experience, polymer-jacketed wires were not used yet. Indeed, Fielder XT wires (Asahi, Japan) – one of the first of its kind – did not obtain the CE mark for commercial use in Europe until 2008 and, to the best of our knowledge, they were never prospectively evaluated neither in bench testing nor in humans, with ELCA. However, the instructions for use of ELCA regarding wire compatibility only mention that a 0.014” (or smaller) guidewire needs to be used to cross the target lesion, and no limitations or caution recommendations are provided for polymer-jacketed wires despite this gap in the evidence.

In the present investigation, including the most used workhorse and CTO-dedicated hydrophilic and polymer-jacketed wires, we sought to examinate the impact of ELCA in terms of wire disruption in the setting of coronary uncrossable lesions. **(Central Illustration)** The main findings can be summarized as follows: 1) In coronary uncrossable lesions ELCA seemed to be a safe technique over hydrophilic wires; 2) The overall incidence of wire disruption after ELCA was significantly higher in polymer-jacketed wires as compared to hydrophilic wires; 3) Higher incidence of “grade 2” disruption was observed in polymer jacketed wires when ELCA was performed with high settings suggesting that further investigation is required in this regard and, until then, ELCA use over polymeric wires must be cautious.

### Incidence and current tools for uncrossable lesions

In the series of Pagnotta and colleagues, where CTO balloon-uncrossable lesions were treated with rotational atherectomy (RA), a microcatheter could not be advanced across the lesions in 22% of the patients and in 4.5% of them it was impossible to advance the Rotawire (Boston Scientific, USA) as well. [19] In the last years ELCA has become a widespread tool and nowadays represents one of the last options in the algorithm for the management of uncrossable lesions, when a microcatheter or a balloon cannot be advanced through the lesion despite a successful distal wiring [12,15]. In CTO interventions, as described by a sub-analysis of the PROGRESS-CTO registry, ELCA was used in up to 21% of the cases given its unique advantages. [14] Above all, in contrast with rotational or orbital atherectomy (OA) devices, which require specific wire (0.009’’ Rotawire for RA and 0.012’’ Viper-wire for OA) for its use, ELCA can be applied over any standard 0.014’’ coronary wire. This can guarantee the passage without the need to exchange the wire through a microcatheter or can even allow a synergistic approach with additional RA/OA use when, after ELCA debulking, a microcatheter for wire exchange can be placed (RASER technique) [20]. With improvement in operator experience and with advances in technology, ELCA has become a widespread tool in coronary intervention, being used in almost 50,000 procedures in USA in the last 30 years. [10,11]

### Coronary wires technology and ELCA

Evidence concerning the safety and the effects of prolonged ELCA exposure over the wire structure is lacking. In recent years many technological iterations in coronary wires have been introduced, ensuring higher performances and lower risk during complex coronary procedures.

According to the type of coating and cover, coronary wires can be classified in hydrophobic, hydrophilic and hydrophilic with polymer-jacketed cover. In polymer-jacketed wires, a polymeric additional layer (cover) is applied over the helical metallic coils, incapsulating them. These polymer layers can be made from a variety of suitable polymers, mostly represented by polyurethane thermoplastic elastomers [21]. In further detail, Teleflex polymer-jacketed wires are covered by an aliphatic polyether-based thermoplastic polyurethane (Tecoflex^®^) and Abbott polymer-jacketed wires are covered by an aromatic polyester-based thermoplastic polyurethane (Pellethane^®^). The detailed characteristics of the polyurethane used for Asahi polymer-jacketed wires were not provided by the company. All these polymers, when exposed to heat start melting at around 190°C (193-226°C for Tecoflex^®^ and 199-205°C for Pellethane^®^). [22] This can be relevant when ELCA is performed, because with each pulse the pressure wave generated by ELCA induces a bubble formation, which ultimately explodes because the heat induced by the process is much higher than the vaporization temperature of water (100°C). The exact temperature of this bubble is very difficult to predict, but it may get significantly closer to the melting temperature of polyurethane, especially for bubbles induced by high ELCA settings or when contrast is also used.

This could have important clinical implications. First, the potential embolization caused by the melting of polymer-jacketed cover of coronary wires could cause distal occlusion of small intramyocardial arteries and generate chronic inflammation with eosinophilic granulocytes as a reaction on the foreign material. [23, 24] Secondly, especially when dealing with uncrossable lesions, a disruption of the wire may often lead to procedure abortion to avoid excessive procedure time, contrast and radiations. In the case shown in **Supp.Fig. 1**, after polymer cover melted, the wire had to be removed and it was confirmed that any balloon or microcatheter could be advanced over the wire beyond the melted segment even outside of the patient. Finally, the polytetrafluoroethylene (PTFE) used in polymer jacketed wires, although is generally safe, under excessive heating can break down and releases molecules as perfluoroisobutene which has high electrophilicity and decomposes rapidly forming various reactive intermediates and fluorophosgene, which then decomposes into carbon dioxide, a radical anion and hydrogen fluoride that might pose a serious health hazard to several human organs. [25,26]

### Practical recommendations

In our in vitro analysis, after ELCA simulation, no wire rupture or de-coiling (“grade 3” event) were observed. However, the coronary artery of our in vitro simulation was filled with saline solution instead of blood or contrast and this could have underestimated the potential amount of wire disruption caused by ELCA. When ELCA is performed with blood, the blood proteins absorb the laser energy creating microbubbles that increase the photomechanical effects at the site of ELCA delivery. This effect is even greater when contrast (either pure or diluted) is simultaneously injected. [27] For this reason, ELCA manufacturers recommend to routinely perform ELCA under continued saline infusion (at a rate of 1 m/sec after a bolus of 5 ml) to avoid interference from microbubble formation, which carries higher risk of coronary complications. [8,28] However, in balloon uncrossable lesions, such as in CTO antegrade intervention with successful wire crossing, antegrade injections are usually avoided to lower the risk of extending coronary dissection. In these scenarios ELCA is usually performed without injecting saline or contrast, but only with coronary blood. In this setting we strongly recommend avoiding the use of ELCA over polymer-jacketed wires since it may lead to higher grade of wire disruption.

When facing challenging lesions polymeric wires represent a cornerstone in the strategies to reach the distal vessel. They often allow to track narrow microchannels or severely calcific plaque thank to the additional layer often made by thermoplastic polyurethane, which guarantee an exceptional lubricity and trackability; they also represent the key device for knuckling in sub- intimal-reentry techniques. In this last scenario if crossing is still challenging after knuckling, the use of ELCA is discouraged according to our findings and exchange for a non-polymeric wire is highly recommended. Manufacturers might consider developing wires with short distal polymeric coating (∼5cm) dedicated for knuckle which might still be safe for the use of ELCA over the non- jacketed segment.

Finally, in the scenario where ELCA remains the only alternative but a polymer-jacketed wire is in place, we highly recommend first trying to exchange the wire for a non-polymeric one and, if impossible or judged too risky, ELCA should be settled at low frequency and pulse fluence (25 pulses/sec and 45 mj/mm2) since most events occurred at higher settings, and strictly avoiding local presence of blood or contrast. (**Figure 2).**

**Figure 2:**
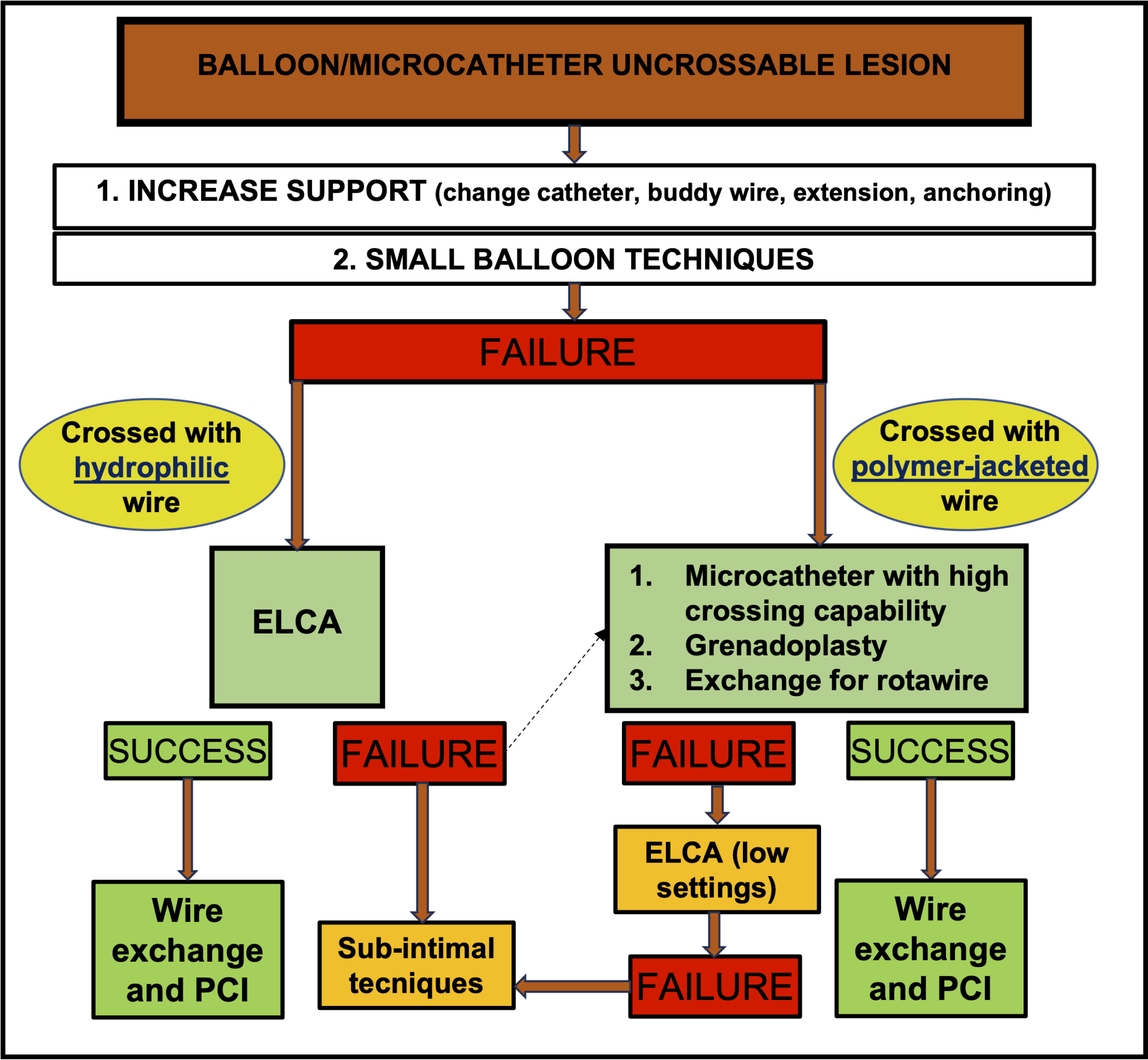
new proposed algorithm, for balloon/microcatheter uncrossable lesions.***Legend:*** new algorithm of balloon/microcatheter uncrossable lesions. The choice of the crossing technique is made according to the type of coronary wire that crossed the lesion. In case of hydrophilic wire ELCA remain a safe and effective option. In case of polymer- jacketed wire, additional attempts to cross a microcatheter are suggested and rotational atherectomy is preferred to ELCA. In these cases, the authors suggest ELCA (along with sub-intimal techniques) as last available option and discourage its use with high settings. Abbreviations: ELCA: excimer laser coronary atherectomy; RA: rotational atherectomy; RASER: a combination of excimer laser coronary atherectomy (ELCA) with rotational atherectomy (RA) (see ref. [20]).

All these precautions are probably no necessary for hydrophilic non-polymeric wires since only minor wire disruptions were observed after ELCA.

## LIMITATIONS

Our analysis has several limitations. First, it represents an in-vitro analysis and, therefore, further in vivo studies are warranted to confirm our findings. Secondly, even if we chose the most common workhorse wires along with the most used specific CTO wires, our findings are limited to the wires that were selected and cannot be extended to all the available coronary wires. Third, as previously discussed, we did not perform our experiment with contrast or blood but only with the “safer” saline solution (as suggested by the ELCA manufacturers), so that further analysis is needed to confirm the safety of ELCA also in these potentially more harmful scenarios.

## CONCLUSIONS

ELCA represent a safe option for coronary balloon uncrossable lesions, especially when used over hydrophilic wires or with low pulse frequency and fluence. The use of ELCA over polymer-jacketed wires should be avoided but, if decision to use is taken, a prudential approach ought to be recommended at the light of our findings, keeping low ELCA settings and always with saline infusion. However, a clinical prospective evaluation of the eventual impact of the use of ELCA over polymer-jacketed wires is mandatory, including the detection of clinical events, the search for circulating toxic molecules, and the imaging evaluation of the impact on microcirculation.

## CLINICAL PERSPECTIVES

**What is known?** ELCA represents one of the last available options in case of balloon uncrossable lesions.

**What is new?** Wire disruption after ELCA is more common for polymer-jacketed wires, especially when high ELCA settings are used.

**What is next?** Further in vivo studies to assess ELCA safety with more potentially harmful settings and to confirm our findings in vivo.

## Data Availability

Data are available for other investigators under request.

## ABBREVIATIONS

CTO: chronic total occlusion
ELCA: excimer laser coronary atherectomy.
OA: orbital atherectomy.
RA: rotational atherectomy.

---

**Central illustration:**
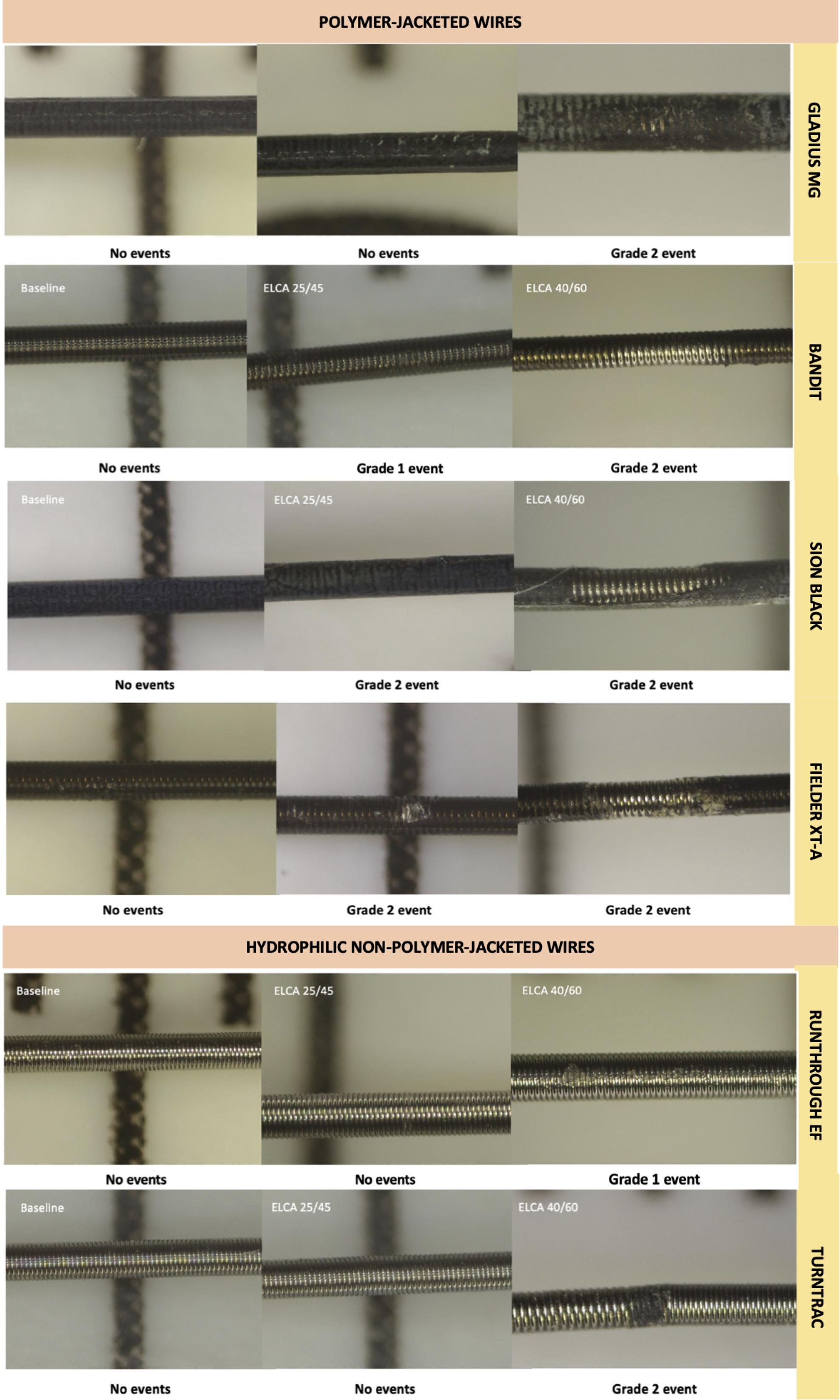
Examples of wire disruption events according to wire type and ELCA settings. ***Legend:*** Examples of various degree of wire disruption are showed in this figure according to the wire type and to the ELCA settings. “ELCA 25/45” indicates 7 pulses of ELCA performed with laser frequency of 25 pulses/min and laser fluence of 45 mj/mm2. “ELCA 40/60” indicates 7 pulses of ELCA performed with laser frequency of 40 pulses/min and laser fluence of 60 mj/mm2. Abbreviations: ELCA: excimer laser coronary atherectomy.

## REFERENCES

1 Elrayes MM, Xenogiannis I, Nikolakopulos I, et al. An algorithmic approach to balloon- uncrossable coronary lesions. Catheter Cardiovasc Interv. 2021;97(6):E817–E825.

2 McQuillan C, Jackson MWP, Brilakis E, et al. Uncrossable and undilatable lesions-A practical approach to optimizing outcomes in PCI. Catheter Cardiovasc Interv. 2021;97(1):121–126.

3 Patel SM, Pokala NR, Menon RV, et al. Prevalence and treatment of “balloon-uncrossable” coronary chronic total occlusions. J Invasive Cardiol. 2015;27(2):78–84.

4 Karacsonyi J, Karmpaliotis D, Alaswad K, et al. Prevalence, indications and management of balloon uncrossable chronic total occlusions: Insights from a contemporary multicenter US registry. Catheter Cardiovasc Interv. 2017;90(1):12–20.

5 Di Mario C, Ramasami N. Techniques to enhance guide catheter sup- port. Catheter Cardiovasc Interv. 2008;72(4):505–512.

6 Kovacic JC, Sharma AB, Roy S, et al. GuideLiner mother-and-child guide catheter extension: a simple adjunctive tool in PCI for balloon uncrossable chronic total occlusions. J Interv Cardiol. 2013;26(4): 343–350.

7 Roy J, Hill J, Spratt JC. The “side-BASE technique”: combined side branch anchor balloon and balloon assisted sub-intimal entry to resolve ambiguous proximal cap chronic total occlusions. Catheter Cardiovasc Interv. 2018;92(1):E15–E19.

8 Tsutsui RS, Sammour Y, Kalra A, et al. Excimer Laser Atherectomy in Percutaneous Coronary Intervention: A Contemporary Review. Cardiovasc Revasc Med. 2021; 25:75–85.

9 Cook SL, Eigler NL, Shefer A, et al. Percutaneous excimer laser coronary angioplasty of lesions not ideal for balloon angioplasty. Circulation. 1991;84:632–43.

10 Koster R, Kahler J, Brockhoff C, et al. Laser coronary angioplasty: history, present and future. Am J Cardiovasc Drugs. 2002;2:197–207.

11 Estella P, Ryan Jr TJ, Landzberg JS, et al. Excimer laser-assisted coronary angioplasty for lesions containing thrombus. J Am Coll Cardiol. 1993;21:1550–6.

12 Karacsonyi J, Armstrong EJ, Truong HTD, et al. Contemporary use of laser during percutaneous coronary interventions: insights from the laser veterans affairs (LAVA) multicenter registry. J Invasive Cardiol. 2018;30:195–201.

13 Ambrosini V, Sorropago G, Laurenzano E, et al. Early outcome of high energy laser (excimer) facilitated coronary angioplasty ON hARD and complex calcified and ballOon- resistant coronary lesions: LEONARDO study. Cardiovasc Revasc Med. 2015;16:141–6.

14 Danek BA, Karatasakis A, Tajti P, et al. Incidence, Treatment, and Outcomes of Coronary Perforation During Chronic Total Occlusion Percutaneous Coronary Intervention. Am J Cardiol. 2017;120(8):1285–1292.

15 Fernandez JP, Hobson AR, McKenzie D, et al. Beyond the balloon: excimer coronary laser atherectomy used alone or in combination with rotational atherectomy in the treatment of chronic total occlusions, non-crossable and non-expansible coronary lesions. EuroIntervention. 2013;9: 243–50.

16 Holmes Jr DR, Forrester JS, Litvack F, et al. Chronic total obstruction and short-term outcome: the Excimer Laser Coronary Angioplasty Registry experience. Mayo Clin Proc. 1993;68:5–10.

17 Topaz O, Ebersole D, Das T, et al. Excimer laser angioplasty in acute myocardial infarction (the CARMEL multicenter trial). Am J Cardiol. 2004;93:694–701.

18. Tai Z., et al. Coronary Atherectomy and Transradial Access Part I of III: Laser Atherectomy. 2016. Vol 24, Issue 8.

19 Pagnotta P, Briguori C, Mango R, et al. Rotational atherectomy in resistant chronic total occlusions. Catheter Cardiovasc Interv. 2010; 76(3):366–371.

20 Egred M. RASER angioplasty. Catheter Cardiovasc Interv. 2012;79: 1009–1012.

21. Richardson MT, et al, Polymer coated guidewires United States Patent (US 6,673,025 B1). 2004.

22 Szycher M. Szycher’s handbook of polyurethanes, second edition. Boca Raton, FL. Taylor & Francis Group. 2013.

23 Grundeken MJ, Li X, Kurpershoek CE, et al. Distal embolization of hydrophilic-coating material from coronary guidewires after percutaneous coronary interventions. Circ Cardiovasc Intervention; 2015;8(2):e001816.

24 Mehta RI, Solis OE, Jahan R, et al. Hydrophilic polymer emboli: an under-recognized iatrogenic cause of ischemia and infarct. Mod Pathol. 2010;23:921– 930.

25 Arroyo CM. The Chemistry of Perfluoroisobutylene (PFIB) with Nitrone and Nitroso Spin Traps: an EPR/Spin Trapping Study. Chem Biol Interact. 1997;105(2):119–29.

26 Lailey AF, Hill L, Lawston IW, Stanton D, Upshall, DG. Protection by Cysteine Esters Against Chemically Induced Pulmonary Oedema. Biochem Pharmacol 1991;42 Suppl:S47- 54.

27 Latib A, Takagi K, Chizzola G, et al. Excimer Laser LEsion modification to expand non- dilatable stents: the ELLEMENT registry. Cardiovasc Revasc Med. 2014;15:8–12.

28 Deckelbaum LI, Natarajan MK, Bittl JA, et al. Effect of intracoronary saline infusion on dissection during excimer laser coronary angioplasty: a randomized trial. The Percutaneous Excimer Laser Coronary Angioplasty (PELCA) Investigators. J. Am Coll Cardiol. 1995;26(5):1264–9.

